# Genetic Distinctions Between Reticular Pseudodrusen and Drusen: Insights from a Genome-Wide Association Study

**DOI:** 10.1101/2024.09.18.24313862

**Authors:** Roy Schwartz, Alasdair N. Warwick, Anthony P. Khawaja, Robert Luben, Hagar Khalid, Sumita Phatak, Mahima Jhingan, Coen de Vente, Philippe Valmaggia, Sandra Liakopoulos, Abraham Olvera-Barrios, Clara I. Sánchez, Catherine Egan, Roberto Bonelli, Adnan Tufail

## Abstract

**Purpose:** To explore genetic determinants specific to reticular pseudodrusen (RPD), and to compare these with genetic associations for drusen.

**Setting:** Participants with RPD, drusen, or controls from the UK Biobank (UKBB), a large, multisite, community-based cohort study.

**Methods:** A previously validated deep learning framework was deployed on 169,370 optical coherence tomography (OCT) volumes from the UKBB to identify cases with RPD and/or drusen and controls without these phenotypes. Cases were included if they were 60 years or older, with at least 5 lesions. Five retina specialists manually validated the cohorts using OCT and color fundus photographs. Quantifications of RPD and drusen derived by the framework were used as variables. Two primary genome-wide association study (GWAS) analyses were performed to explore potential genetic associations with number of RPD and drusen within ‘pure’ cases, where only RPD or drusen were present in either eye. A candidate approach was furthermore adopted to assess 46 previously known AMD loci. Secondary GWAS were undertaken for number of RPD and drusen in mixed cases, as well as binary case-control analyses for pure RPD and pure drusen. Genome-wide significance was defined as p<5e-8.

**Results:** A total of 1,787 participants were identified and analysed, including 1,037 controls, 361 pure drusen, 66 pure RPD and 323 mixed cases. The primary pure RPD GWAS yielded four genome-wide significant loci: rs11200630/*ARMS2-HTRA1* (p=1.9e-09), rs79641866/*PARD3B* on chromosome 2 (p=1.3e-08), rs143184903/*ITPR1* on chromosome 3 (p=8.1e-09), and rs76377757/*SLN* on chromosome 11 (p=4.3e-08). The latter three are all uncommon variants (minor allele frequency <5%). A significant association at *CFH* was also observed adopting a candidate approach (p=1.8e-04).

Two loci reached genome-wide significance for the primary pure drusen GWAS: rs10801555/*CFH* on chromosome 1 (p=6.0e-33) and rs61871744/*ARMS2-HTRA1* on chromosome 10 (p=4.2e-20). For the mixed RPD and drusen secondary analyses, lead variants at both the *CFH* and *ARMS2-HTRA1* loci reached genome-wide significance, with *C2-CFB-SKIV2L* additionally associated for mixed drusen alone. Findings from the binary case-control GWAS for drusen mirrored those of the primary drusen analysis, however no variants reached genome-wide significance in the case-control RPD GWAS.

**Conclusions:** Our findings indicate a clear association between the *ARMS2*-*HTRA1* locus with higher RPD load. Although the association at the *CFH* locus did not reach genome-wide significance, we observed a suggestive link. We furthermore identified three novel associations that are unique to RPD, albeit for uncommon genetic variants. These findings were only observable when quantifying RPD load as a continuous trait, which increased the study’s statistical power. Further studies with larger sample sizes are now required to explore the relative contributions of *AMRS2-HTRA1* and *CFH* to RPD development, and to explore the validity of these newly presented RPD-specific genetic associations.

## Introduction

Age-related macular degeneration (AMD) is the leading cause of severe, irreversible vision loss in Europe and North America. It predominantly affects older individuals and can lead to central vision loss by affecting the macula, an area of the retina that is important for detailed visual tasks such as reading and facial recognition. Traditionally, the presence of drusen - acellular, lipid-rich deposits beneath the retinal pigment epithelium (RPE), characterizes AMD.^1,2^ As a result of advancements in retinal imaging, reticular pseudodrusen (RPD), also known as subretinal drusenoid deposits (when detected on optical coherence tomography (OCT) scanning), have been identified as another manifestation of the AMD spectrum.

Several lines of evidence highlight the distinctions between drusen and RPD. Anatomically, drusen are situated beneath the retinal pigment epithelium (RPE), whereas RPD are located above it.^3^ Additionally, their biochemical compositions vary: drusen comprise both esterified and non-esterified cholesterol, while RPD predominantly contain unesterified cholesterol.^4^ In addition, a recent study showed that RPD are abundant in lysolipids, that are not found in drusen.^5^ Numerous studies indicate that the presence of RPD in eyes with AMD increases the risk of rapid progression to late AMD stages, including geographic atrophy (GA) and neovascularisation (mostly type 3 macular neovascularization (MNV)) compared to drusen without RPD.^6,7^ Our research has also discovered a correlation between a higher load of RPD and stroke,^8^ and other studies have suggested links between RPD and coronary artery disease, as well as lower levels of high-density lipoprotein (HDL).^9^ In contrast, large studies have not consistently found these cardiovascular associations with drusen-associated AMD.^10–12^ Moreover, meta-analyses have not confirmed a link between AMD and cardiovascular diseases or stroke.^13,14^ RPD can also appear in conditions where drusen are not part of the main phenotype, such as pseudoxanthoma elasticum^15^, Late-Onset Retinal Degeneration,^16^ and vitamin A deficiency.^17^

Approximately 46-71% of AMD variation is estimated to be heritable.^18^ The largest single genome-wide association study (GWAS) to date in AMD identified 52 independently associated common and rare variants distributed across 34 loci.^19^ Chief among them is complement factor H (CFH) on chromosome 1, identified as a genetic contributor to AMD in the earliest studies.^20–24^ Another major susceptibility locus is the *ARMS2*-*HTRA1* region on chromosome 10.^25–27^ In combination, these two loci are estimated to account for more than half the genetic predisposition to AMD.28,29

The genetic basis of RPD is not as well characterized. Previous studies examining RPD genetics have been small and focused only on known AMD-associated risk loci. The most consistently associated loci for RPD are *ARMS2*-*HTRA1* followed by *CFH*, although conflicting effect directions have been reported for the latter (Supplementary Table 1).^9,30–43^

Given the differences between RPD and drusen, we theorized that RPD and drusen arise from both shared and distinct biological pathways. Identifying pathways that are unique to RPD could aid in developing targeted therapies. To test this theory, we conducted a GWAS using retinal imaging data from the UK Biobank (UKBB) study, leveraging a validated AI algorithm to identify and quantify drusen and RPD on this large dataset.^44^ Our aim was to explore potential genetic associations that are specific to RPD and compare them with associations found for drusen.

## Methods

### UK Biobank

The UKBB study is a large, multisite, community-based cohort study designed to advance the prevention, detection, and treatment of a wide range of severe and potentially fatal diseases. This study contains data from 500,000 predominantly self-described white ethnicity volunteers, aged between 40 and 69, recruited across the United Kingdom between 2006 and 2010.

Eligibility for the study was extended to all UK residents within the specified age group who were registered with the National Health Service (NHS) and resided within a 25-mile radius of any of the 22 assessment centers. The study was approved by the North West Multi-centre Research Ethics Committee (REC reference number: 06/MRE08/65, UKBB project ID 60078), adhering to the principles of the Declaration of Helsinki (www.ukbiobank.ac.uk). Participants who withdrew their consent were excluded from the study.

A sub-cohort of 67,687 participants underwent OCT and color fundus photography (CFP) imaging at six centers (Sheffield, Liverpool, Hounslow, Croydon, Birmingham, and Swansea). Images were captured using the Topcon 3D OCT 1000 Mark II (Topcon, Japan) under mesopic conditions without pupil dilation using the 3-dimensional macular volume scan (512 horizontal A-scans/B-scan; 128 B-scans in a 6 × 6-mm raster pattern).

### Study population

The research utilized a deep learning framework to distinguish between subjects with RPD, drusen, and control groups within the UKBB dataset. The specifics of this framework have been outlined in a prior publication.^44^ In brief, the process involved an image classifier that filtered out ungradable OCT B-scan volumes. This was followed by a deep ensemble model that detected out-of-distribution data, further eliminating ungradable volumes. Once these volumes were removed, another classifier evaluated the gradable B-scan volumes to categorize subjects into groups with drusen, RPD, or control groups. The final step of the process employed a semantic segmentation model to quantify the number of RPD and drusen within each B-scan. Of note, although the UKBB contains both CFP and OCT scans for participants with retinal imaging, we chose to analyze only OCT B-scans within the framework given findings from previous studies showing their higher sensitivity for RPD compared with CFP.^45,46^ However, CFPs were used in human validation of the framework’s output, as detailed below.

The AI framework was applied to the entire UKBB OCT dataset to identify subjects with drusen, RPD, and a control group without either of these phenotypes. The study included subjects identified by the AI framework based on the following criteria: a. Age 60 years or older for all groups; b. At least five instances of RPD or drusen when these phenotypes were present. This requirement was set to rule out cases with minimal lesions, which could be due to natural variation. c. For subjects with drusen, the size of the drusen had to surpass 63 microns in at least one eye. This ensured the exclusion of subjects with only “drupelets” (which in population studies are common features and not shown to increase the risk of progression to late AMD),^1^ while including subjects with at least early AMD as per the Beckman classification.^1^ To accurately measure the size of drusen, we determined the maximum diameter of drusen in any direction (Feret diameter) on the en face plane.

Two retina specialists (H.K., R.S.) graded the cases identified by the AI framework for the presence of RPD, drusen, or both. Multimodal imaging, including OCT and CFP, was used for validation. Cases with advanced AMD (GA or MNV) were excluded as significant changes in the outer retina from neovascular AMD or GA could affect the accuracy of lesion quantification. Any disagreement between graders was adjudicated by a senior grader (A.T.).

Four retina specialists (A.T., S.P., M.J., R.S.) validated the control group through multimodal imaging assessments, including OCT volumes and CFP, to ensure the absence of drusen and RPD. CFPs complemented OCT scans to detect any drusen or RPD outside the field of view of OCT scans. Subjects with ungradable CFPs or with imaging artifacts that would prevent a confirmation of a subject’s control status (e.g., shadowing on CFPs, artifacts affecting the outer retina on OCT) were excluded.

Following grader validation of cases, the number of RPD and drusen identified by the semantic segmentation model on all B-scan in the volumes of both eyes for each patient was summed, representing the total lesion count per patient.

### Genome-wide association study

Full details for genotyping and imputation in the UKBB cohort have been described previously.^47^ In brief, genotype calling was performed using two arrays: the UK BiLEVE Axiom array (∼50,000 participants) and the UK Biobank Axiom array (∼450,000 participants). Marker positions are in GRCh37 coordinates. There were 805,426 markers available in the released data after quality control. Genotype imputation was then performed using a combined Haplotype Reference Consortium and UK10K reference panel, expanding the number of testable variants to ∼96 million.

The following exclusions were applied for sample quality control: individuals with relatedness corresponding to third-degree relatives or closer, excess of missing genotypes or more heterozygosity than expected. The GWAS analysis was furthermore restricted to individuals of European ethnicity only due to the lack of power for other genetic backgrounds. The following exclusions were applied for variant-level quality control: call rate <95%, Hardy-Weinberg equilibrium p<1 x 10^-6^, posterior call probability <0.9, INFO score <0.9 and minor allele frequency <0.01.

GWAS analyses were performed to assess potential genetic associations with (i) pure RPD (defined as cases with RPD in at least one eye but without drusen in either eye) and (ii) pure drusen (defined as cases with drusen in at least one eye but without RPD in either eye). For each analysis, the study cohort included participants with only the phenotype of interest and control participants, as described earlier. The primary analyses were based on the number of RPD and drusen; the count data, which included zero values for control participants, was standardized by adding one to each value prior to log transforming to achieve normal distributions. Secondary analyses were also performed with the additional inclusion of mixed cases (cases with simultaneous presence of drusen and RPD) for number of RPD and number of drusen, as well as binary case-control analyses for pure RPD and pure drusen compared against controls. All GWAS analyses were conducted using a generalised linear mixed model, adjusting for age, sex and the first ten principal components. Correction for multiple testing was performed by applying a genome-wide significance threshold of p < 5x10^-8^.

### Candidate approach

For the two primary GWAS analyses (number of pure RPD and pure drusen), a candidate approach was furthermore adopted based on 34 advanced AMD-associated loci previously reported by Fritsche et al^19^ and 12 early AMD loci from Winkler et al.^48,49^. Loci were considered to pass experiment-wise significance at p < 0.001 (0.05/46).

### Number of Pure RPD Post-GWAS analysis

Summary statistics results were investigated using FUMA.^50^ The SNP2GENE protocol as used to perform positional and expression quantitative trait locus (eQTL) gene mapping/prioritisation.^50^ SNPs in high linkage disequilibrium (LD; r2>0.6) with any independent lead variant were positionally mapped to genes located within 10kb. Variants were also mapped to a set of prioritized genes within 1Mb if associated with the expression of those genes in retina (reported in EyeGEx^51^) and blood (reported in eQTLGen^50^).

Novel loci (not previously associated with AMD) were further evaluated by deriving credible sets and annotating these to investigate for variants with functional relevance, as well as assessing for colocalization between the RPD signals with gene expression. A Bayesian fine-mapping approach was used to generate 95% credible sets of putative causal variants for genome-wide significant loci. Variants within 500 kb of each lead variant were included, applying a prior probability of 10^-4^ for each variant. Fine-mapped variants were retained if they were significantly associated with RPD at p<0.05 and in linkage disequilibrium with the lead variant at r2>0.6. The OpenTargets Genetics database was then queried for variant annotation.^52,53^ Colocalization between novel RPD-associated loci with gene expression was assessed for all genes within 500kb of each lead variant, for all tissues within the GTEx v8 project with at least one significant *cis*-eQTL for those genes.^54^ Overlapping variants within 500kb of each lead variant were included. Results were deemed supportive of colocalization if the maximum probability for PP.H4 (both traits are associated and share a single causal variant) was greater than 0.75.

Previously reported AMD-associated loci (that either reached genome-wide significance or passed the experiment-wise significance threshold with the candidate approach) were assessed for colocalization with AMD, using summary statistics from FinnGenn (H7_AMD, release 10)^55,56^ Variants within 500kb of each lead variant from our RPD GWAS were included.

### Statistical Software

GWAS analyses were performed using REGENIE software to account for relatedness across individuals.^57^ Genomic inflation factor and heritability estimates were calculated using the LDSC tool^58^ and pre-calculated LD scores for European ancestry.^59^ All other statistical analyses were performed in R (R for GNU macOS, Version 4.2.0, The R Foundation for Statistical Computing, Vienna, Austria).^60^ The R package coloc was used for fine-mapping and colocalization analyses.^61–63^ Other R packages used included targets^64^, tarchetypes^65^, tidyverse^66^, workflowr^67^, flextable^68^, gtsummary^69^ and knitr^70^.

## Results

A total of 1,787 European participants met the study inclusion and quality control criteria, including 1,037 controls with no drusen or RPD, 66 participants with RPD but no drusen (‘pure RPD’), 361 participants with drusen but no RPD (‘pure drusen’), and 323 participants with a mixture of drusen and RPD. The median (interquartile range) number of lesions in the two pure groups was 91 (39-299) RPD and 201 (94-397) drusen, while for the mixed group there were 73 (31-208) RPD and 233 (96-516) drusen. Demographic characteristics are summarized in Table 1.

**Table 1.** Demographic characteristics.

|  | Controls<br>N = 1,037 | RPD<br>N = 66 | Drusen<br>N = 361 | Mixed RPD/drusen<br>N = 323 |
| --- | --- | --- | --- | --- |
| Age | 62.6 (60.7, 65.2) | 65.6 (63.2, 68.0) | 64.5 (61.6, 66.6) | 64.2 (60.8, 66.5) |
| Female | 570 (55%) | 34 (52%) | 216 (60%) | 150 (46%) |

Age presented as median (interquartile range); sex presented as N (%) female.

### Primary GWAS for number of pure RPD and pure drusen

The primary GWAS analyses yielded 4 loci reaching genome-wide significance for number of pure RPD (*ARMS2-HTRA1*, *PARD3B*, *ITPR1* and *SLN*) and 2 loci for number of pure drusen (*CFH* and *ARMS2-HTRA1*), summarized in **Table 2** and **Figure 2**. We did not observe any inflation of the GWAS statistics (genomic inflation factor = 1.005 and 1.008, RPD and drusen respectively). The estimated SNP heritability for number of RPD was 0.35 (0.41), while that for number of drusen was 0.08 (0.39). Genetic correlation analyses were not possible due to the limited SNP heritability.

**Figure 1.**
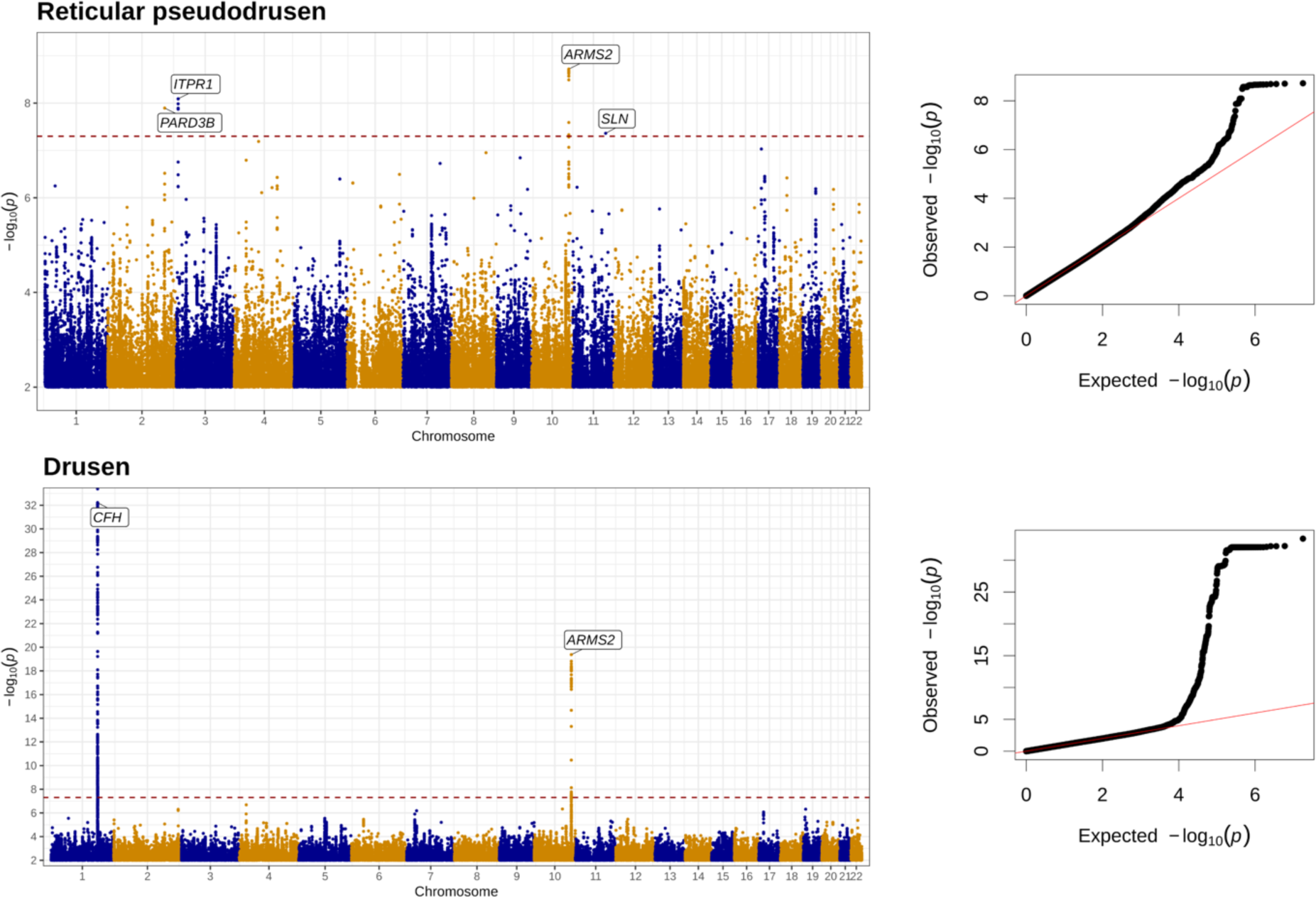
Manhattan and quantile-quantile plots of GWAS results for number of pure RPD (in the absence of drusen) and number of pure drusen (in the absence of RPD)

**Table 2.** Genome-wide significant associations with number of pure reticular pseudodrusen and pure drusen.

|  | **Rs identifier** | **chr:pos[hg19]** | **EA/OA (EAF)** | **Beta (95% CI)** | **P** | **Nearest gene** |
| --- | --- | --- | --- | --- | --- | --- |
| **Reticular pseudodrusen** |  |  |  |  |  |  |
|  | rs79641866 | 2:206022440 | T/C (0.03) | 0.39 (0.25; 0.53) | 1.3e-08 | *PARD3B* |
|  | rs143184903 | 3:4692390 | G/A (0.01) | 0.64 (0.42; 0.87) | 8.1e-09 | *ITPR1* |
|  | rs11200630 | 10:124209684 | C/T (0.21) | 0.16 (0.11; 0.21) | 1.9e-09 | *ARMS2* |
|  | rs76377757 | 11:107598700 | T/C (0.01) | 0.58 (0.37; 0.79) | 4.3e-08 | *SLN* |
| **Drusen** |  |  |  |  |  |  |
|  | rs10801555 | 1:196660261 | A/G (0.46) | 0.35 (0.29; 0.41) | 6.0e-33 | *CFH* |
|  | rs61871744 | 10:124203787 | C/T (0.24) | 0.3 (0.23; 0.37) | 4.2e-20 | *ARMS2* |

Both loci for drusen (*CFH* and *ARMS2-HTRA*) are well known to be associated with AMD, while among the four found for RPD only the *AMRS2*-*HTRA1* locus has previously been reported in association with AMD. The lead variants associated with RPD at *PARD3B* (rs79641866) and *ITPR1* (rs143184903) are both intronic variants, whereas rs76377757 lies in an intergenic region ∼8000 base pairs downstream of the *SLN* locus (Supplementary Figure S1). It should be noted that the minor allele frequencies for all three novel RPD-associated loci are uncommon (MAF < 0.05).

We then adopted a candidate-based approach to look up 34 previously reported AMD-associated variants for AMD and 12 variants for early AMD.^19,48,49^ This yielded three additional loci for number of drusen (*C9*, *C3* and *CFI*) and one additional locus for number of RPD (*CFH*) that met the experiment-wise significance threshold (p < 0.05/46; Supplementary Table 2), all of which have previously been associated with advanced AMD.

### Number of pure RPD post-GWAS analysis

Gene mapping did not prioritise any additional genes at the novel *PARD3B*, *ITPR1* and *SLN* loci. We calculated 95% credible sets at each of the three loci and investigated for any overlap with functional regulatory regions but did not find evidence of this. Colocalization analyses also did not show any evidence for colocalization between these RPD signals with gene expression for any of the tissues with available data from the GTEx database.

Colocalization analyses for previously known AMD loci (*CFH* and *AMRS2-HTRA1*) did, however, show strong evidence for colocalization with AMD GWAS statistics from Finngen. The estimated posterior probabilities for a shared causal variant between number of pure RPD with AMD were 79% and 100% for *CFH* and *ARMS2-HTRA1,* respectively (Supplementary Table 3).

### Secondary GWAS analyses

We subsequently performed GWAS for number of RPD and number of drusen, this time including cases where both RPD and drusen were present (mixed cases). Lead variants at *CFH* and *ARMS2-HTRA1* loci reached genome-wide significance for both analyses, with additional associations near *CFH* at *CFHR4* for mixed RPD, and both *CFHR4* and *CFHR5* for mixed drusen. Furthermore, the *C2-CFB-SKIV2*L locus was genome-wide significant for mixed drusen only (Supplementary Table 4, Supplementary Figure S2).

Finally, GWAS analyses were undertaken using binary case-control definitions for pure drusen and pure RPD. Results from the binary pure drusen analysis mirrored those from the primary GWAS for number of pure drusen, with lead variants at both CFH and *AMRS2-HTRA1* loci reaching genome-wide significance (Supplementary Figure S3, Supplementary Table 5). For the binary pure RPD GWAS however, no variants reached genome-wide significance (Supplementary Figure S3).

## Discussion

We present findings from the first reported genome-wide association studies for RPD, confirming an association between *ARMS2*-*HTRA1*, a known AMD locus, with higher RPD load. In contrast to drusen, *CFH* was not associated with number of RPD at genome-wide significance. However, an association with *CFH* was observed at a less stringent experiment-wise significance threshold and colocalization analyses indicated a shared causal variant for RPD and AMD at this locus. We furthermore identified three novel associations with uncommon lead variants at *PARD3B*, *ITPR1* and *SLN*, although these should be interpreted with caution given the limited power of our study.

The associations between *CFH* and *ARMS2*-*HTRA1* with AMD are well established, representing the earliest genome-wide significant findings from any GWAS.^20–27^ In combination, these two loci are estimated to account for more than half the genetic predisposition to AMD.^28,29^ It is striking how the significance of the *CFH* locus, the most significant locus for drusen, is far less significant for RPD. While this is likely in part due to the difference in power, the *ARMS2-HTRA1* locus which is less significant for drusen remains genome-wide significant for RPD. This may suggest that *ARMS2-HTRA1* has a more important role for RPD than *CFH*, but further functional work would be necessary to test this hypothesis. Previous RPD studies have consistently demonstrated an association with *ARMS2*-*HTRA1*, but reported mixed results for *CFH* (Supplementary Table 1).^71^ It is important to recognize the limitations of previous studies, which may explain the discrepancy between previous and current findings, as well as among different previous studies (Supplementary Table 6).

A critical challenge in identifying the genetic origin of RPD relates to the lack of sensitive imaging modalities for large AMD cohorts. Numerous genetic studies primarily relied on CFP to ascertain the presence of RPD. However, CFP exhibits reduced sensitivity compared to other imaging techniques.^38,39,41,45,46,72^ Consequently, this may lead to an underestimation of RPD prevalence. Our approach involved their identification and quantification on OCT B-scans, which have been shown to have the highest sensitivity (alongside near-infrared (NIR) imaging) for the detection of RPD. Furthermore, the literature reveals a lack of consensus regarding the number of distinct RPD necessary to categorize an eye as RPD-positive. Methodologies range from requiring a single RPD to a minimum of five. The process of retinal aging leads to physiological alterations in the outer retina, which may resemble RPD or drusen in small quantities. Therefore, a more judicious approach in the grading of RPD might entail stipulating a specific number of lesions for a definitive diagnosis. However, only three previous studies had this as a requirement. The detection of RPD in cases of advanced AMD is further complicated due to the extensive retinal damage caused by AMD. Several studies have included such cases. In our study, such cases were excluded.

As an additional limitation, previous research has primarily focused on a select set of known AMD-associated risk loci. This approach overlooks the potential genetic contribution from other AMD risk loci, as well as variants not directly associated with AMD, which could be involved in diverse pathophysiological processes. Given the distinct differences between RPD and drusen, it is plausible that independent and novel pathological pathways may be instrumental in their formation. Therefore, we adopted a hypothesis-free approach to identify genetic associations with RPD.

Similarly, the majority of previous studies were limited by their inclusion of RPD solely in the presence of drusen-associated AMD. Only six of these studies considered pure RPD as part of their analysis, but they were underpowered due to the small number of pure RPD cases (the largest number of pure RPD was 30).^36^ Our study has the largest number of pure RPD cases reported in the literature. Another potential difference that could explain our findings is the relatively young age of our pure RPD cohort (mean 65.6 years, compared with an overall mean of 77.5 years in previous studies). The occurrence of RPD at an earlier age may correspond more closely with inheritance, whereas late-onset findings may be more influenced by environmental factors or lifestyle choices that accumulate over time, as well as age-related changes.

The methodology used in our primary analyses differs from the commonly used case-control GWAS study design. Instead, the load of RPD and drusen were used to identify genetic associations, thereby overcoming the low statistical power due to the paucity of cases. Our results indicate that this methodology is valid, given the findings previously known to be associated with these pathologies and found using more traditional techniques. While we could not identify any variants associated with RPD reaching genome-wide significance in a case-control analysis, these associations were exposed when examining the load of RPD.

Our research provides insights into the pathophysiology of RPD. The *ARMS2* gene is expressed in the ellipsoid region of the photoreceptor inner segments,^73^ near the subretinal space, where RPD have been observed using OCT and histological analysis, unlike the sub-RPE location of drusen.^3,74^ While the exact role of ARMS2 remains unknown, it is speculated to be linked to mitochondrial function,^75^ extracellular matrix turnover,^76,77^ or both. These processes might contribute to photoreceptor damage and cell death, leading to the accumulation of debris identified as RPD.

The close linkage of *HTRA1* with *ARMS2* also may explain the association we observed. The *HTRA1* gene encodes a secreted enzyme that is believed to regulate cell growth and break down various extracellular matrix proteins.^78,79^ The AMD risk haplotype is associated with changes in the levels of *HTRA1* expression.^80,81^ Therefore, the association between *HTRA1* and RPD might be due to *HTRA1*-mediated increased breakdown of the extracellular matrix, resulting in debris accumulation that appears as RPD.

We also identified three novel loci, not reported previously in association with RPD. It is notable that all three novel variants are uncommon, with a MAF of less than 0.05 and therefore should be interpreted with caution pending validation in larger studies and external validation cohorts. PARD3B (Par-3 Family Cell Polarity Regulator Beta), is predicted to enable phosphatidylinositol binding activity. Additionally, it is predicted to be involved in the establishment of cell polarity and maintenance of epithelial cell apical/basal polarity (strictly apical in many epithelia) and regulated by TGFb signalling.^82^ Given this role, if PARD3B is involved in the regulation of membrane localization and the organization of cellular components in retinal cells, it could potentially affect where deposits form relative to the RPE, thereby explaining the differential location of RPD compared to drusen.

ITPR1 (Inositol 1,4,5-Trisphosphate Receptor Type 1) encodes an intracellular receptor for inositol 1,4,5-trisphosphate. Upon stimulation by inositol 1,4,5-trisphosphate, this receptor mediates calcium release from the endoplasmic reticulum. ITPR1 plays a role in endoplasmic reticulum stress-induced apoptosis. Mutations in this gene cause spinocerebellar ataxia type 15, spinocerebellar ataxia type 29, as well as Gillespie Syndrome, which may involve iris hypoplasia (verify and add Adnan reference) [Ref RefSeq, Nov 2009]. ^83,84^

The *SLN* gene encodes sarcolipin, a small proteolipid that regulates several sarcoplasmic reticulum Ca(2+)-ATPases.. Sarcoplasmic reticulum Ca(2+)-ATPases are transmembrane proteins that catalyze the ATP-dependent transport of Ca(2+) from the cytosol into the lumen of the sarcoplasmic reticulum in muscle cells. Sarcolipin interacts with Ca(2+)-ATPases and reduces the accumulation of Ca(2+) in the sarcoplasmic reticulum without affecting the rate of ATP hydrolysis.^85^

Early onset drusen maculopathy has been associated with both common and rare, potentially deleterious, genetic variants (with MAF< at 1% level). In a genetic association study, Breuk et al. identified rare variants predominantly involving the complement and lipid metabolism pathways – which were not previously noted in association studies of patients with conventional age of onset AMD.^86^ Similarly, patients included in our cohort of pure RPD were much younger than previously studied cohorts of RPD patients, who mostly have the combined RPD and drusen phenotype. Although these findings should be replicated in a larger cohort of RPD patients, to elucidate their contribution to RPD, the novel genetic associations found may give us insights into pathways and mechanism of RPD formation.

In summary, we present the first reported GWAS for RPD, finding a genome-wide significant association with ARMS2-HTRA1, and potentially novel associations with PARD3B, ITPR1 and SLN. An association with CFH was also observed, though at a lower significance threshold, and this locus colocalized with AMD. Larger studies are needed to clarify the roles of ARMS2-HTRA1 and CFH in RPD development and to validate these newly identified RPD-specific genetic associations. Future research should aim to quantify RPD burden to enhance the statistical power of these studies.

## Data Availability

Data produced in the present study are available upon reasonable request to the authors

## Acknowledgements

Sponsored by the EURETINA Retinal Medicine Clinical Research Grant.

R.S., C.E., and A.T. received a proportion of their financial support from the UK Department of Health through an award made by the National Institute for Health Research to Moorfields Eye Hospital NHS Foundation Trust and UCL Institute of Ophthalmology for a Biomedical Research Centre for Ophthalmology.

A.P.K is supported by a UK Research and Innovation Future Leaders Fellowship, an Alcon Research Institute Young Investigator Award and a Lister Institute for Preventive Medicine Award. This research was supported by the NIHR Biomedical Research Centre at Moorfields Eye Hospital and the UCL Institute of Ophthalmology.

This work was supported in part by the Said Foundation

We want to acknowledge the participants and investigators of the FinnGen study.

For the purpose of open access, the author has applied a Creative Commons Attribution (CC BY) licence to any Author Accepted Manuscript version arising

