## Supplementary material for "Genetic Distinctions Between Reticular Pseudodrusen and Drusen: Insights from a Genome-Wide Association Study"

### Supplementary Tables

**Supplementary Table 1 – Genetic associations of reticular pseudodrusen identified in previous studies**

| Source | CFH Y402H† | ARMS2 | HTRA1 | CFH I62V† | C3 | C2/CFB‡ | VEGFA | LIPC | SKIV2L |
|---|---|---|---|---|---|---|---|---|---|
| Klein et al., (Beaver Dam Eye Study), 2008 [30] | ↑ | -- | -- | -- | -- | -- | -- | -- | -- |
| Smith et al., 2011 [31] | ↓ | ↑ | -- | -- | -- | -- | -- | -- | -- |
| Ueda-Arakawa et al., 2013 [32] | 0 | ↑ | -- | 0 | -- | -- | -- | -- | -- |
| Puche et al., 2013 [33] | ↑ | ↑ | ↑ | -- | ↑ | -- | -- | -- | -- |
| Joachim et al., (Blue Mountain Eye Study), 2014 [34] | ↑ | ↑ | -- | -- | -- | -- | -- | -- | -- |
| Yoneyama et al., 2014 [35] | -- | ↑ | -- | 0 | -- | -- | -- | -- | -- |
| Boddu et al., 2014 [36] | 0 | 0 | -- | -- | -- | -- | -- | -- | -- |

Continued on next page

| Source | CFH Y402H† | ARMS2 | HTRA1 | CFH I62V† | C3 | C2/CFB‡ | VEGFA | LIPC | SKIV2L |
|---|---|---|---|---|---|---|---|---|---|
| Finger et al. (Melbourne Collaborative Cohort Study), 2016 [37] | ↑ | ↑ | ↑ | ↑ | -- | -- | -- | -- | -- |
| Buitendijk et al., (Rotterdam Study), 2016 [38] | ↑ | ↑ | -- | ↓ | ↑ | 0 | ↑ | 0 | ↓ |
| Wu et al., 2016 [39] | 0 | ↑ | -- | -- | -- | -- | -- | -- | -- |
| Lin et al., (CATT trials), 2018 [40] | 0 | ↑ | ↑ | 0 | 0 | 0 | -- | 0 | -- |
| Domalpally et al., 2019 [41] | 0 | ↑ | -- | -- | 0 | 0 | 0 | -- | -- |
| Dutheil et al., (Alienor Study), 2020 [42] | ↑ | ↑ | -- | -- | 0 | 0 | -- | ↑ | -- |

Continued on next page

| Source | CFH Y402H† | ARMS2 | HTRA1 | CFH I62V† | C3 | C2/CFB‡ | VEGFA | LIPC | SKIV2L |
|---|---|---|---|---|---|---|---|---|---|
| Altay et al. (EUGENDA Study), 2021 [43] | -- | ↑ | -- | ↓ | -- | -- | -- | -- | ↓ |
| Thomson et al., 2022 [9] | 0 | ↑ | -- | -- | 0 | 0 | -- | 0 | -- |

↑ - increased risk, ↓ - decreased risk, 0 no risk, -- SNP not investigated in study

Gene variants were only added to the list if they appeared in more than one study. Otherwise a description of findings is in the text.

†SNP rs800292 refers to CFH I62V and SNP s1061170 to CFH Y402H

‡C2 and CFB are in linkage disequilibrium. They are shown together or separately in different studies

**Supplementary Table 2. Candidate approach to identify genetic associations with number of (i) reticular pseudodrusen and (ii) drusen.**

| | Rs identifier | chr:pos[hg19] | EA/OA (EAF) | Beta (95% CI) | P | Nearest gene | Advanced or early AMD locus |
|---|---|---|---|---|---|---|---|
| **Reticular pseudodrusen** | | | | | | | |
| | rs10922109 | 1:196704632 | C/A (0.61) | 0.09 (0.04; 0.13) | 1.8e-04 | *CFH* | Advanced |
| | rs3750846 | 10:124215565 | C/T (0.21) | 0.16 (0.1; 0.21) | 2.1e-09 | *ARMS2-HTRA1* | Advanced |
| **Drusen** | | | | | | | |
| | rs10922109 | 1:196704632 | C/A (0.66) | 0.32 (0.26; 0.39) | 5.6e-25 | *CFH* | Advanced |
| | rs10033900 | 4:110659067 | T/C (0.48) | 0.1 (0.04; 0.16) | 4.5e-04 | *CFI* | Advanced |
| | rs62358361 | 5:39327888 | T/G (0.02) | 0.52 (0.28; 0.76) | 1.5e-05 | *C9* | Advanced |
| | rs3750846 | 10:124215565 | C/T (0.25) | 0.29 (0.22; 0.35) | 8.9e-19 | *ARMS2-HTRA1* | Advanced |
| | rs2230199 | 19:6718387 | C/G (0.22) | 0.15 (0.08; 0.22) | 2.5e-05 | *C3* | Advanced |

**Supplementary Table 3. Colocalization results between GWAS findings for number of pure reticular pseudodrusen and age-related macular degeneration (Finngen) at the *CFH* and *ARMS2-HTRA1* loci. Maximum probabilities are highlighted in bold.**

| Gene symbol | Lead RSID | PP.H0 | PP.H1 | PP.H2 | PP.H3 | PP.H4 |
|---|---|---|---|---|---|---|
| *CFH* | rs1061170 | 7.2e-269 | 1.3e-268 | 7.4e-02 | 0.13 | **0.79** |
| *ARMS2-HTRA1* | rs11200630 | - | - | 1.8e-06 | 4.2e-03 | **1.00** |

**Supplementary Table 4. Genome-wide significant associations with number of mixed reticular pseudodrusen and mixed drusen.**

| | Rs identifier | chr:pos[hg19] | EA/OA (EAF) | Beta (95% CI) | P | Nearest gene |
|---|---|---|---|---|---|---|
| **Reticular pseudodrusen** | | | | | | |
| | rs9970075 | 1:196673430 | T/G (0.49) | 0.38 (0.31; 0.45) | 2.3e-26 | *CFH* |
| | 1:196884765_CA_C | 1:196884765 | CA/C (0.91) | 0.39 (0.26; 0.53) | 3.8e-09 | *CFHR4* |
| | rs201396317 | 10:124213674 | A/C (0.24) | 0.32 (0.24; 0.4) | 9.9e-16 | *ARMS2* |
| **Drusen** | | | | | | |
| | rs10922102 | 1:196668287 | C/T (0.49) | 0.45 (0.39; 0.51) | 6.8e-51 | *CFH* |
| | rs141073581 | 1:196851265 | A/G (0.96) | 0.48 (0.31; 0.65) | 2.1e-08 | *CFHR4* |
| | rs9427662 | 1:196946775 | T/C (0.92) | 0.35 (0.23; 0.47) | 3.4e-09 | *CFHR5* |
| | rs429608 | 6:31930462 | G/A (0.87) | 0.27 (0.18; 0.36) | 9.3e-10 | *SKIV2L* |
| | rs61871744 | 10:124203787 | C/T (0.27) | 0.34 (0.28; 0.41) | 1.2e-26 | *ARMS2* |

**Supplementary Table 5. Genome-wide significant associations with presence or absence of pure drusen.**

| | Rs identifier | chr:pos[hg19] | EA/OA (EAF) | OR (95% CI) | P | Nearest gene |
|---|---|---|---|---|---|---|
| **Drusen** | | | | | | |
| | 1:196661838_TTTTC_T | 1:196661838 | TTTTC/T (0.44) | 3.32 (2.7; 4.08) | 9.3e-34 | *CFH* |
| | rs61871744 | 10:124203787 | C/T (0.24) | 2.54 (2.06; 3.13) | 2.4e-19 | *ARMS2* |

#### Supplementary Table 6 – Characterization of previous genetic studies for reticular pseudodrusen

| Source | Imaging modalities | | | | | | | Minimal number of RPD required | Late AMD only | Genes explored | Pure RPD | Mean age |
|---|---|---|---|---|---|---|---|---|---|---|---|---|
| | CFP | NIR | SD-OCT | FAF | RF | FA | ICG | | | | | |
| Klein et al., (Beaver Dam Eye Study), 2008 [30] | x | | | | | | | NA | No | AMD partial | 0 | 61.8 |
| Smith et al., 2011 [31] | x | x | | x | | | | NA | No | AMD partial | 0 | 83 |
| Ueda-Arakawa et al., 2013 [32] | x | x | x | x | | x | x | 5 | Yes | AMD partial | 0 | 73.9 |
| Puche et al., 2013 [33] | | x | x | | | | | NA | No | AMD partial | 0 | 77.4 |
| Joachim et al., (Blue Mountain Eye Study), 2014 [34] | x | | | | | | | NA | No | AMD partial | Unclear | 69.5 |

Continued on next page

| Source | Imaging modalities | | | | | | | Minimal number of RPD required | Late AMD only | Genes explored | Pure RPD | Mean age |
|---|---|---|---|---|---|---|---|---|---|---|---|---|
| | CFP | NIR | SD-OCT | FAF | RF | FA | ICG | | | | | |
| Yoneyama et al., 2014 [35] | x | x | x | x | x | | | NA | Yes | AMD partial | 0 | 74.9 |
| Boddu et al., 2014 [36] | | x | x | | | | | NA | No | AMD partial | 30 | 87* |
| Finger et al. (Melbourne Collaborative Cohort Study), 2016 [37] | x | | | | | | | NA | No | AMD partial | 6 | 76 |
| Buitendijk et al., (Rotterdam Study), 2016 [38] | x | x | | | | | | NA | No | AMD partial | 14 | 82.1 |
| Wu et al., 2016 [39] | | x | x | | | | | 5 | No | AMD partial | 0 | 76.2 |

Continued on next page

| Source | Imaging modalities | | | | | | | Minimal number of RPD required | Late AMD only | Genes explored | Pure RPD | Mean age |
|---|---|---|---|---|---|---|---|---|---|---|---|---|
| | CFP | NIR | SD-OCT | FAF | RF | FA | ICG | | | | | |
| Lin et al., (CATT trials), 2018 [40] | x | | | | x | x | | NA | No | AMD partial | 0 | 78 |
| Domalpally et al., 2019 [41] | x | | | x | | | | 5 | No | AMD | 0 | 79 |
| Dutheil et al., (Alienor Study), 2020 [42] | x | x | x | x | | | | NA | No | AMD partial | 22 | 81.9 |
| Altay et al. (EUGENDA Study), 2021 [43] | | x | x | | | | | NA | No | AMD | 9 | 80.7 |
| Thomson et al., 2022 [9] | | x | x | x | | | | NA | No | AMD partial | 29 | 81.3 |

AMD = age-related macular degeneration, CFP = Colour fundus photography, FA = Fluorescein angiography, FAF = Fundus autofluorescence, ICG = Indocyanine green, NIR = Near infrared photography, RF = Red-free, RPD = reticular pseudodrusen, SD-OCT = Spectral domain optical coherence tomography

* - Median age. Mean age was not provided.

AMD partial = only some of known AMD-associated genetic loci were explored

### Supplementary figures

**Supplementary figure S1. Locus zoom plots for lead variants from the genome-wide association study on number of reticular pseudodrusen.**

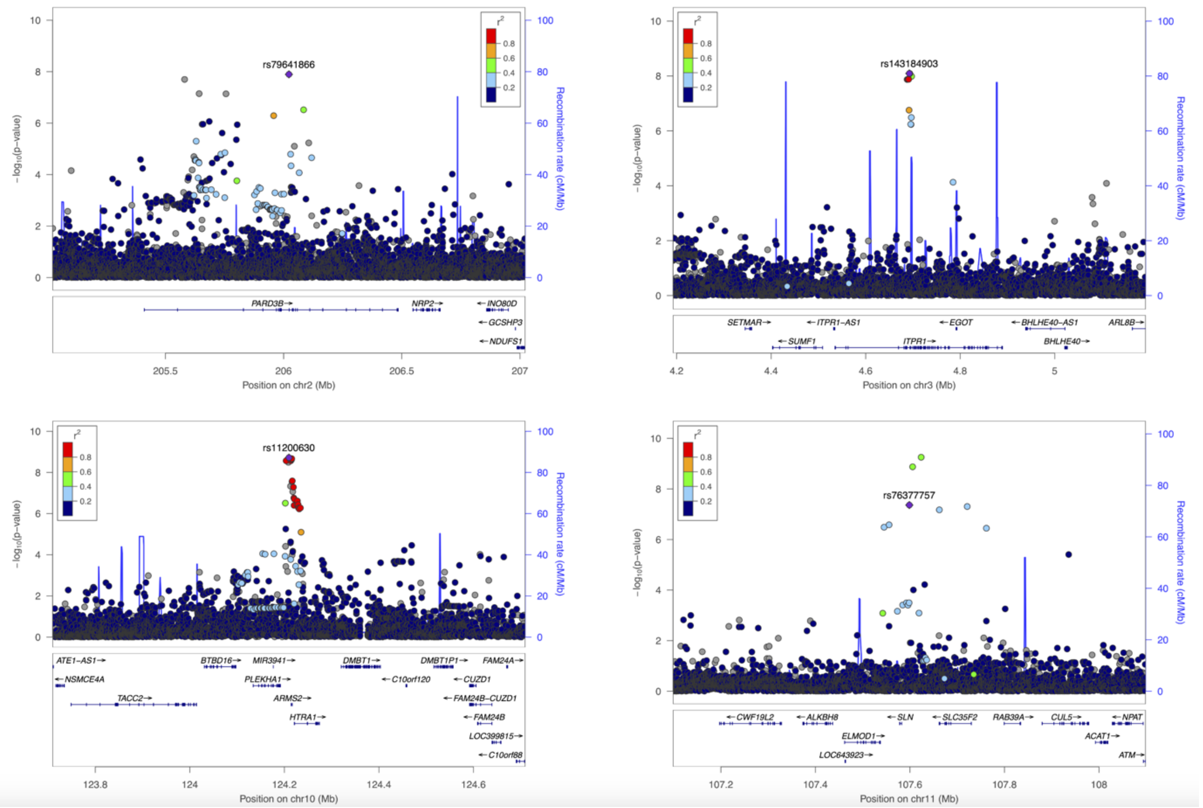

**Supplementary figure S2. Manhattan and quantile-quantile plots of continuous GWAS results for mixed RPD with or without drusen) and mixed drusen with or without RPD).**

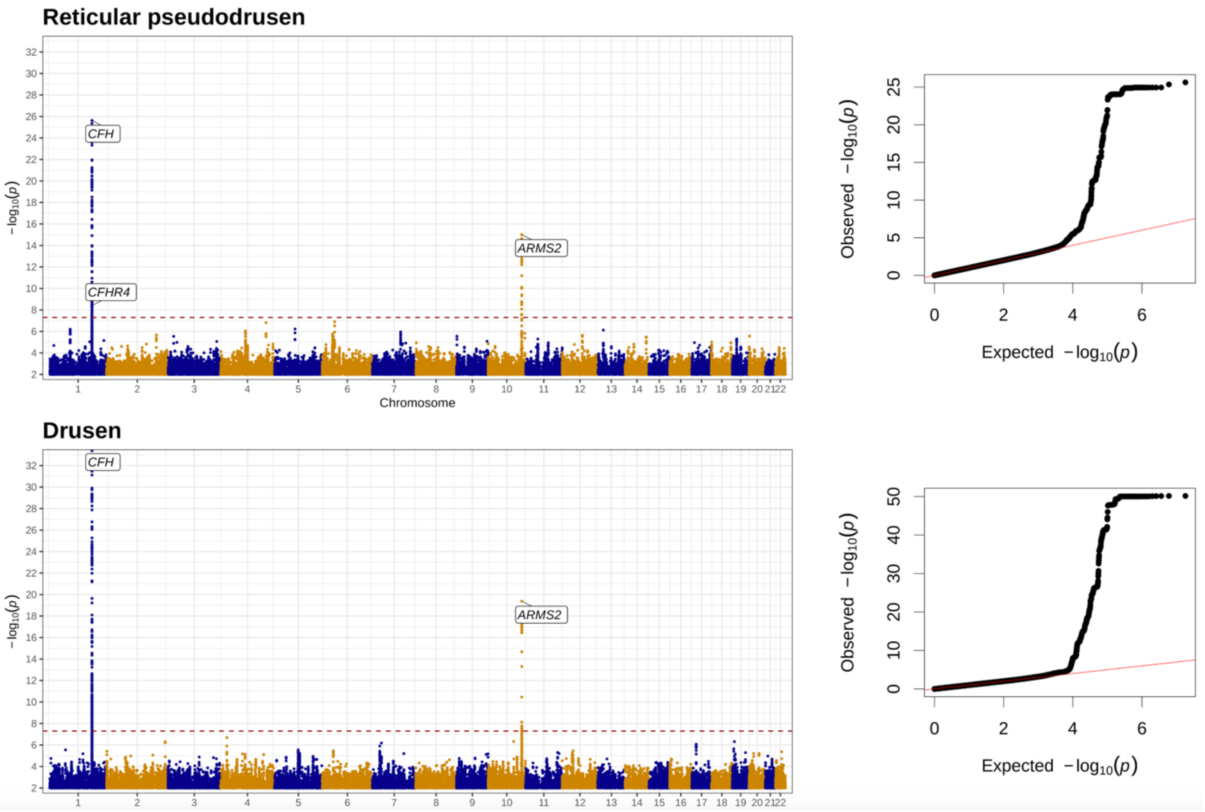

**Supplementary figure S3. Manhattan and quantile-quantile plots of case-control GWAS results for pure RPD (in the absence of drusen) and pure drusen (in the absence of RPD)**

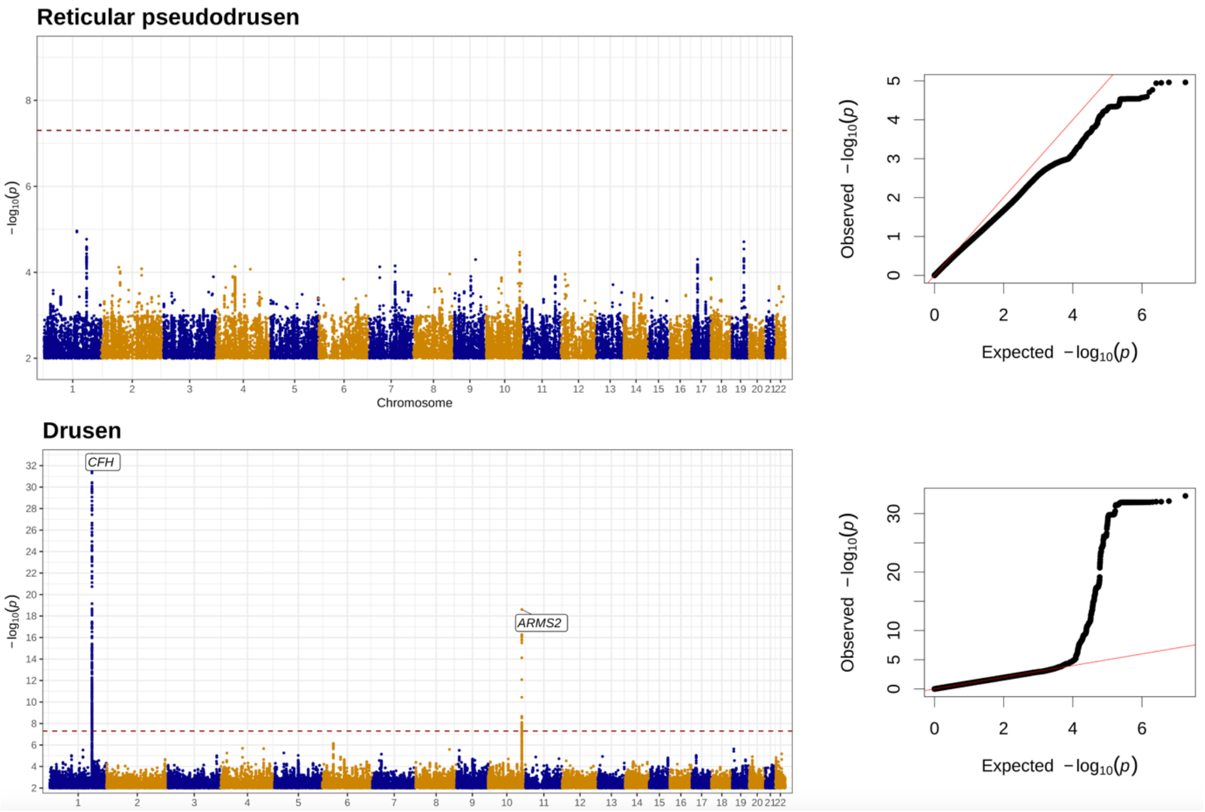
